# Diagnostic accuracy and safety of coaxial core-needle biopsy (CNB) system in Oncology patients treated in a specialist cancer centre with prospective validation within clinical trial data

**DOI:** 10.1101/2020.04.17.20065458

**Authors:** Khurum Khan, Reyes Gonzalez exposito, David Cunningham, Dow-Mu Koh, Andrew Woolston, Louise Barber, Beatrice Griffins, Kyriakos Kouvelakis, Vanessa Calamai, Monia Bali, Nasir Khan, Annette Bryant, Claire Saffery, Charles Dearman, Ruwaida Begum, Sheela Rao, Naureen Starling, David Watkins, Ian Chau, Chiara Braconi, Nicola Valeri, Marco Gerlinger, Nicos Fotiadis

## Abstract

**Background:** Image guided tissue biopsies are critically important in diagnosis and management of cancer patients. High yield samples are also vital for biomarker and resistance mechanism discovery through molecular/genomic analyses.

**Patients and methods:** All consecutive patients who underwent plugged image-guided biopsy at Royal Marsden from June 2013 until September 2016 were included in the analysis. In second step, a second cohort of patients prospectively treated within two clinical trials (PROSPECT-C and R), were assessed for the DNA yield from biopsies assessed for complex genomic analysis.

**Results:** A total of 522 plugged core biopsies were performed in 457 patients [52% men; median age 63 years (range 17-93)]. Histological diagnosis was achieved in 501/522 (96%) of performed biopsies. Age, gender, modality, metastatic site and seniority of the interventionist were not found to be significant factors associated with odds of failure on a logistic regression. Seventeen (3.3%) were admitted due to biopsy-related complications; 9, 3, 2, 1, 1,1 were admitted for grade I/II pain control, sepsis, vasovagal syncope, thrombosis, haematuria and deranged liver functions respectively; 2 patients with right upper quadrant pain after liver biopsy were found to have radiologically confirmed subcapsular haematoma requiring conservative treatment. One patient (0.2%) developed grade III haemorrhage following biopsy of a gastric GIST tumour. Overall molecular analysis was successful in 89% (197/222 biopsies). Prospective validation in 62 biopsies gave success rates of 92.06% and 79.03% for DNA extraction of >1microgram and tumour content of >20% respectively.

**Conclusion:** The probability of diagnostic success for complex molecular analysis is increased with plugged large co-axial needle biopsy technique, which also minimises complications and reduces hospital stay. High yield DNA acquisition allows genomic molecular characterisation for personalised medicine.

**Statement of significance:** Cancer diagnosis and personalised management is largely dependent on safe acquisition of tumour tissue required for histological diagnosis, and sometimes genomic characterisation. This poses significant challenge to treating physicians, when deliberating risk-benefit ratio of invasive procedures, especially within the context of clinical trials. In this largest examination of safety and efficacy of biopsies in more than 500 patients, we show that diagnostic success for complex molecular analysis is increased with CNB technique that minimises complications and reduces hospital stay. Moreover, we provide validation of our findings with a group of patients treated within prospective clinical trials.

## INTRODUCTION

Whilst cancer management and treatment options have significantly improved during the last few years, our knowledge and understanding about mechanisms of response and/or resistance to anti-cancer therapies remain relatively sparse. To date, this relative lack of understanding is partially due to difficulties in accessing prospectively collected tissue and blood samples from systemic anti-cancer therapy (SACT) -resistant tumours.

Image guided tissue biopsies are not just important in establishing an accurate histo-pathological diagnosis and standard cancer management; high yield samples are also vital in understanding the molecular and genomic characteristics of tumours. Genomic analyses on tumour samples broadly fall into two categories including: 1) targeted approaches investigating a limited number of genes that are known to influence clinical decision making and 2) whole exome or genome sequencing frequently adopted in exploratory research studies to learn about new mechanisms of response or resistance to SACT [1, 2]. Conventional formalin fixed paraffin embedded (FFPE) samples obtained during diagnostic procedures may not be sufficient for such analyses to be realised. For instance, the data from The Cancer Genome Atlas (TCGA) studies showed that fresh frozen material from primary tumour resection specimens was associated with a tumour content of 60% [3]. Moreover, using FFPE DNA for large-scale genomic studies may demonstrate mutations that have occurred as a result of the fixation process, which makes it difficult to distinguish real tumour variants from these fixation artefacts. Furthermore, low quality fragmented DNA can fail quality control in the pre-analytical stage impairing success rates.

Whilst a number of retrospective studies have demonstrated the safety and accuracy of diagnostic biopsies [4-6], data interpretation from such studies has often been hampered by small numbers, the lack of information on yield for molecular/genomic characterisation of tumours, and the lack of prospective validation. At Royal Marsden (RM) we have been using coaxial core-needle biopsy (CNB) system and a pre-formed gelatine sponge sealing device to conduct solid organ core biopsies in order to minimise the number of passes and reduce the risk of complications respectively. We present here the largest dataset demonstrating the safety and accuracy of this approach. Moreover, we took the opportunity to utilise a cohort of patients from two prospective clinical trials to validate tumour yields from biopsies in these translational studies.

## METHODS

### Study design

All consecutive patients who underwent plugged image guided biopsy at RM from June 2013 until September 2016 were included in the analysis. Data including gender, age, primary tumour, biopsy site, needle gauge, interval between biopsy and discharge, incidence of complications and biopsy success were collected. The study was approved by the local institutional review board (Ethics Committee) at the Royal Marsden NHS Foundation Trust. The two trials in the validation cohort including *PROSPECT-C* (clinical trials.gov number [NCT02994888],[2]) and *PROSPECT-R* (clinical trials.gov number [NCT03010722],)[1] were reviewed by the National Research Ethics Committee (NRES) in the UK.

### Biopsy Technique

The biopsies were performed by a Consultant Interventional Radiologist (IR) or an IR fellow under supervision. Ultrasound and CT guidance was used based on the location of the lesion. Conscious sedation was administered along with local anaesthesia, when required, to maximize co-operation and improve patient experience. A 15G or 17G co-axial needle was inserted under direct image guidance at the edge of the lesion and 2-6 cores were obtained with a 16G or 18G automatic core biopsy needle (*True-Core II, Argon Medical Devices, Frisco, Texas, USA*) respectively. Different areas inside the lesion were sampled by changing the angle and position of the co-axial needle. After the samples were collected, one to four preformed 16G or 18G gelatin foam pledgets (*Hunter biopsy sealing device, Vascular solutions Inc, Minneapolis, Minessota, USA*) were deployed through the co-axial along the tract of the needle to facilitate haemostasis. The gelatin resorbs completely within 12 weeks.

### Validation cohort

In the second step, a validation cohort of patients prospectively treated within two clinical trials was used to assess the DNA yield utilised for genomic analysis. The two trials included: *PROSPECT-C* (clinical trials.gov number [NCT02994888],[2]) and *PROSPECT-R* (clinical trials.gov number [NCT03010722],)[1]; phase II, open label, non-randomised studies of anti-epidermal growth factor receptor (EGFR) monoclonal antibodies and regorafenib in patients with *RAS* wild type and *RAS* mutant refractory metastatic colorectal cancer (CRC) respectively. All participants in both studies were required to have mandatory pre-treatment biopsies (6 cores), biopsies at partial response in PROSPECT-C and stable disease at 2 months in PROSPECT-R (6 cores) and at the time of progression (6-12 cores from two suitable progressing metastatic sites).

### Prospective tissue collection procedures

Fresh frozen and FFPE tissue samples were obtained and plasma collection was conducted as per the study protocols at the clinically relevant defined time points. 16-gauge core biopsy was used to collect 3 or 4 fresh biopsy specimens and 1 or 2 specimens fixed in formalin and paraffin embedded. Within the trials, approximately 25% of the total length of a core was detached for primary culture and remaining ∼75% of the core was snap frozen and used for genomic analysis. One core was transported to establish tumour derived organoids and targeted panel validation [7]. One core was used for genomic analysis after being placed into cryovials and immediately snap frozen in liquid nitrogen. The remaining two cores were placed straight into formalin, and embedded in paraffin wax. Primary morphological and immunohistochemical analysis was performed by the histo-pathologist on the FFPE specimen, for confirmation of diagnosis. The samples were then stored in the GI & Lymphoma Research Bank of the RM, anonymised by trial number and time point.

### Tissue sample processing

Biopsy cores were snap frozen in liquid nitrogen at the time of collection. Genomic (g)DNA and mRNA were co-extracted from cores using the Qiagen All-Prep kit. DNA was also isolated from whole blood samples using the Qiagen QIAamp DNA Blood Mini kit.

### Whole exome sequencing

A minimum of 500 nanograms of gDNA was prepared for WES using the Agilent SureSelect Human All Exon v5 capture library, according to the manufacturers’ protocol. The resulting libraries were sequenced to a mean depth of 100x using paired end 100 reads on an Illumina HiSeq 2500. High quality reads were aligned to the NCBI reference genome (hg19) using BWA (v0.7.12), and SAMtools (v0.1.19) to remove duplicates. Tumour content was estimated based on the CNVkit (v0.8.1) copy number profile.

### Sanger Sequencing

For patients with a known tumour variant, PCR was performed on 20 nanograms of gDNA using M13F/R-tailed mutation specific primers (Life Technologies; Supplementary table 2) and Q5 High-Fidelity 2x Master Mix (NEB) on an Eppendorf Mastercycler Nexus GSX1. Primer specific annealing temperatures for Q5 polymerase were established using the NEB online Tm calculator. PCR products were cleaned using Qiaquick PCR purification kit (Qiagen) and 15 nanograms DNA was submitted for M13F and M13R sequencing using the Mix2Seq service (Eurofins Genomics). Ab1 traces were visualised and compared to the reference sequence using ApE software (https://openwetware.org/mediawiki/index.php?title=ApE_-_A_Plasmid_Editor_(software_review)&oldid=753142). Sample tumour content was estimated from the relative abundance of wild-type and variant peaks (**Supplementary Table 1**).

### Statistical design

The success rate of biopsies was determined by the ability to perform standard molecular testing on tissue specimens, and safety determined by frequency of complications and extended hospital stay. Encrypted data was collected in a password protected Excel file and statistical analysis performed usingSTATA13. Chi-squared-analysis was undertaken to identify baseline characteristics that provided independent association with failure and success rates.

## RESULTS

### Overall safety of image guided biopsies and cox regression analysis

A total of 522 tissue biopsies were performed in 457 patients [48% men; median age 63 years (range 23-86)] (**Supplementary Table 2**). Two, three and four biopsies were obtained from 51 (11.2%), 13 (2.8 %), and 1 (0.2%) patients, respectively, at different time points as part of clinical trial protocols. Histological diagnosis was achieved in 501/522 (96%) of performed biopsies. Same-day discharge was achieved for 444 (85.1%) procedures as outpatients, 35 (6.7%) and 17 (3.3%) had planned inpatient and elective procedures respectively and 8 (1.5 %) patients were kept in for overnight observation after a late evening procedure. Seventeen (3.3%) were admitted with the following biopsy-related complications: grade I/II pain control (9), sepsis (1), vasovagal syncope (2), thrombosis (1), haematuria (1) and deranged liver functions (1). Two patients with right upper quadrant pain had radiologically confirmed subcapsular haematoma requiring conservative treatment. One patient (0.2%) developed grade III haemorrhage requiring transfusion of two units of packed red blood cells following biopsy of a gastric GIST. In 21/522 biopsies diagnosis was not achieved due to sampling error during needle placement. These were small lesions not well visualised with ultrasound and CT and normal tissue adjacent to the lesion was consequently biopsied. When patients were divided into two groups including those who underwent “liver biopsy” (n=284 biopsies from 231 patients) and all other biopsies except liver, i.e. “others” (n=238 biopsies from 228 patients). Success rates of 95.02% and 98.32% were observed in the two groups respectively (**Supplementary Tables 3A and B**).

#### Chi-squared tests to assess covariates of failure

Results from chi-squared tests showed that the covariates of age category at earliest biopsy date, gender, modality of image guidance, metastatic site and seniority of the interventionist were not associated with the occurrence of failure. Association of site of biopsy (others vs. liver) however showed a significant trend in favour of others organs vs. liver, although the difference was not found to be numerically and clinically of significant impact (p=0.053). Patients who had biopsy within clinical trials (n=163 biopsies) vs. those who underwent routine clinical diagnostic biopsies (n=338), showed a success rate of 98.79% and 95.48% respectively. Chi-squared test demonstrated significance in favour of patients treated within clinical trials (p=0.07) (**Supplementary Table 4**).

### Validated genomic testing in patients with metastatic colorectal cancer (mCRC)

Given that mCRC patients underwent genomic profiling for clinically actionable mutations such as *KRAS, NRAS*, and *BRAF* analysis routinely with a clinically validated COBAS panel, we rationalized separating this cohort from the remaining patients. Of the total 144 patients with mCRC, 17 repeat charts and 38 patients who were referred from other hospitals were excluded. Of the remaining 89 patients, 2 (2.25%) had a failed molecular analysis due to insufficient DNA extraction-29 (32.58%), 6 (6.74%) and 3 (3.37%) were found to have *KRAS* exon 2-4, *NRAS* exons 2-4 and *BRAF v600* mutations respectively. Moreover, 36 patients were tested for *TP53* and *PIK3CA* mutation; 26 (72.22%) and 5 (13.89%) were found to have these mutations respectively. These results are largely consistent with previously published literature (**Supplementary excel sheet data**).

### gDNA extraction from biopsy: a cohort of PROSPECT-C and R studies

DNA was extracted from 62 biopsies taken from our prospective PROSPECT-C and R trials, and in 65% of cases; sufficient gDNA for WES was achieved from a single core. Two or three tissue cores were needed to yield sufficient DNA for 27% and 8% of the biopsy time points respectively (**Supplementary Figures 1**). When required, utilising all available tissue cores allowed gDNA extraction rate of 100%. Tumour content was determined for 62 biopsies (75.61%) in the analysed cohort and was estimated as greater than 20% in 79.03% of cases (**Table 1**).

**Table 1:**
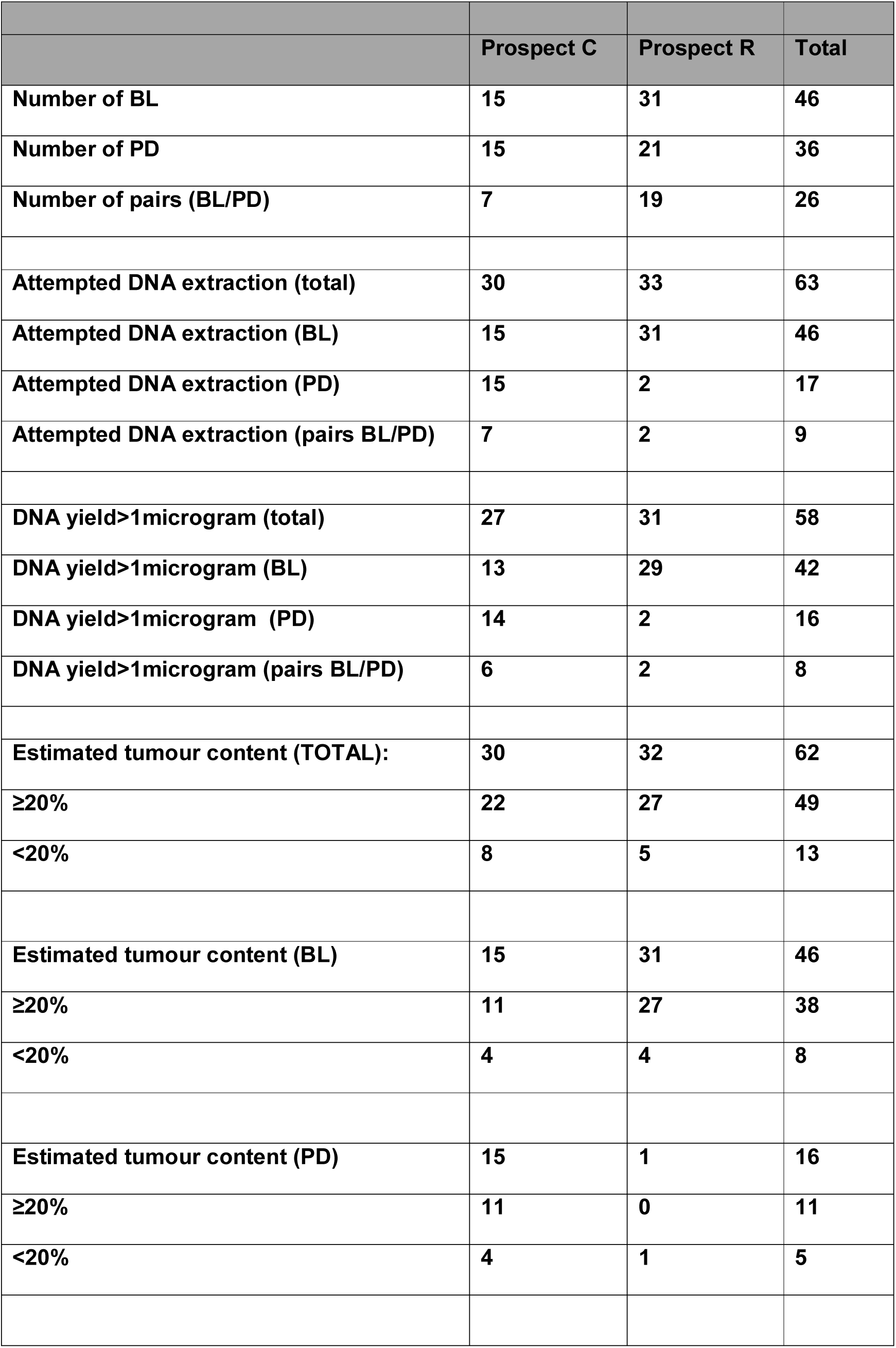
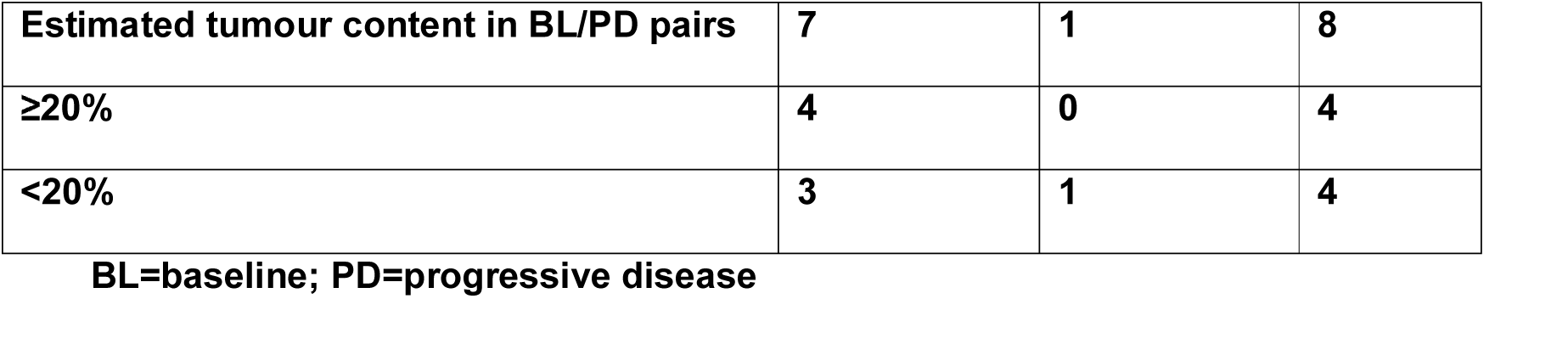
DNA extraction and estimated tumour content

### Assessment of biopsy tumour content

All patients entering the PROSPECT trials were tested for *KRAS*/NRAS mutations in the archival tumour biopsy by standard COBAS methodology as this precluded entry into PROSPECT-C study [2]. As a result, all patients entering the PROSPECT-R trial had a catalogued *KRAS*/*NRAS* variant that could be used to investigate the tumour content of the respective biopsy samples [1]. Mutation profiles for *KRAS/NRAS* have previously been shown to be highly concordant between samples from the same colorectal tumour [8]. We therefore estimated the cancer cell content of biopsy samples using Sanger sequencing to detect the likely truncal *KRAS/NRAS* mutations identified previously by clinical sequencing assays. Samples were scored according to the following criteria: “high” tumour content if the variant base was detected at an intensity exceeding or equal to the wild type base; “medium” if the variant base was detected at more than 25% of the intensity of the wild type base; “low” if the variant base was clearly detected above background but at less than 25% of the intensity of the wild type base; “not detected” if the variant base could only be detected within the background noise or not at all (**Table 2 and Figure 3**). Further cores were extracted and sequenced if the first had low or no detectable tumour content (**Table 2**). In five cases, an additional core had medium tumour content where the first tested core was low/not detectable tumour content. Out of the 49 samples tested, 39 were scored as medium or high tumour content (**Table 2**).

**Table 2:** Sample tumour content estimated from Sanger Sequencing

|  | **Prospect R** |
| --- | --- |
| **Number of samples sequenced:** | 49 |
| **Number of cores sequenced per sample:** |  |
| **1 core:** | 35 |
| **2 cores:** | 10 |
| **3 cores:** | 4 |
| **Number of high tumour content cores:** | 12 |
| **Number of medium tumour content cores:** | 27 |
| **Number of low tumour content cores:** | 6 |
| **Number of cores with no detectable tumour content:** | 22 |

**Figure 1:**
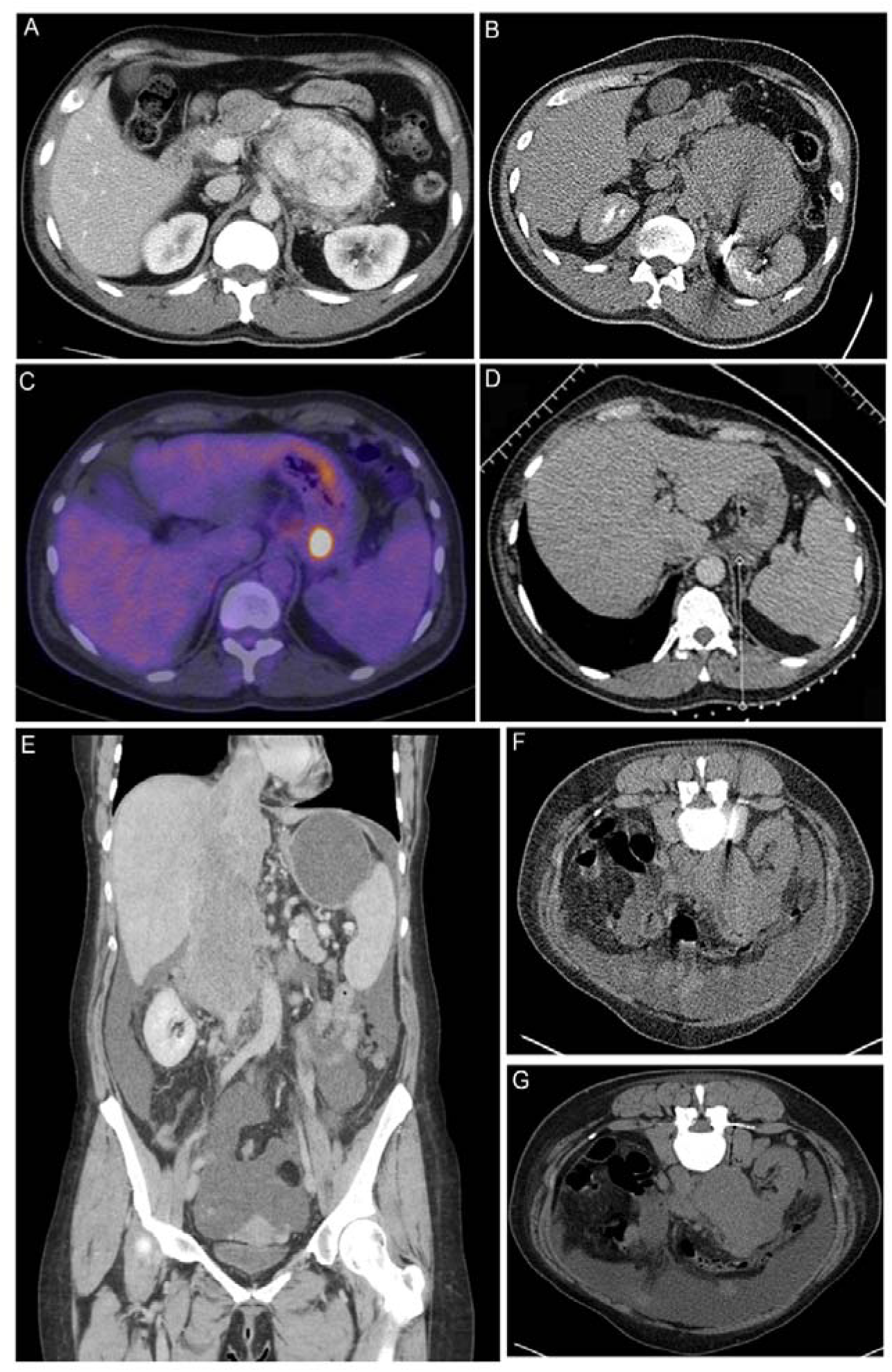
Biopsy examples of patients within the study. **A**) A Computed tomography (CT) of a patient with a highly vascular retroperitoneal mass thought to be too high risk to biopsy at the local hospital. Surgery was also considered to be high risk of R1/R2 resection and a CT guided biopsy was recommended at our MDT. **B**) Biopsy was performed with a 15G/16G co-axial needle. The tract was plugged with16G Hunter plugs and there were no complications. The biopsy showed an Inflammatory Myofibroblastic Tumour which responded well on steroids and an operation was avoided. **C**) PET/CT of a 57yr old patient with relapsed Hodgkin’s Lymphoma after 6 cycles of ABVD chemotherapy. There was response in all sites of disease with the exception of a plaque of tissue behind the fundus of the stomach which appear FDG-avid on PET scan. A decision at the MDT was made to biopsy the lesion in order to exclude transformation of lymphoma. **D**) The 17G co-axial needle was placed medial to the left adrenal and above the splenic vessels adjacent to the lesion. Three cores were taken and the tract was plugged. There were no complications. The biopsy showed Hodgkin’s Lymphoma which responded well to systemic therapy and consolidation RT. **E**) Coronal CT images of a 48yr old patient with a large tumour of the inferior vena cava (IVC) extending from the level of the renal veins to the right atrium. Occluded hepatic veins and ascites can be seen on the scan. **F**) The lesion was biopsied with a 15G/16G co-axial needle. **G**) The tract was plugged with three gelfoam pledgets. A diagnosis of leiomyosarcoma of the IVC was made and procedure had no complications.

**Figure 2:**
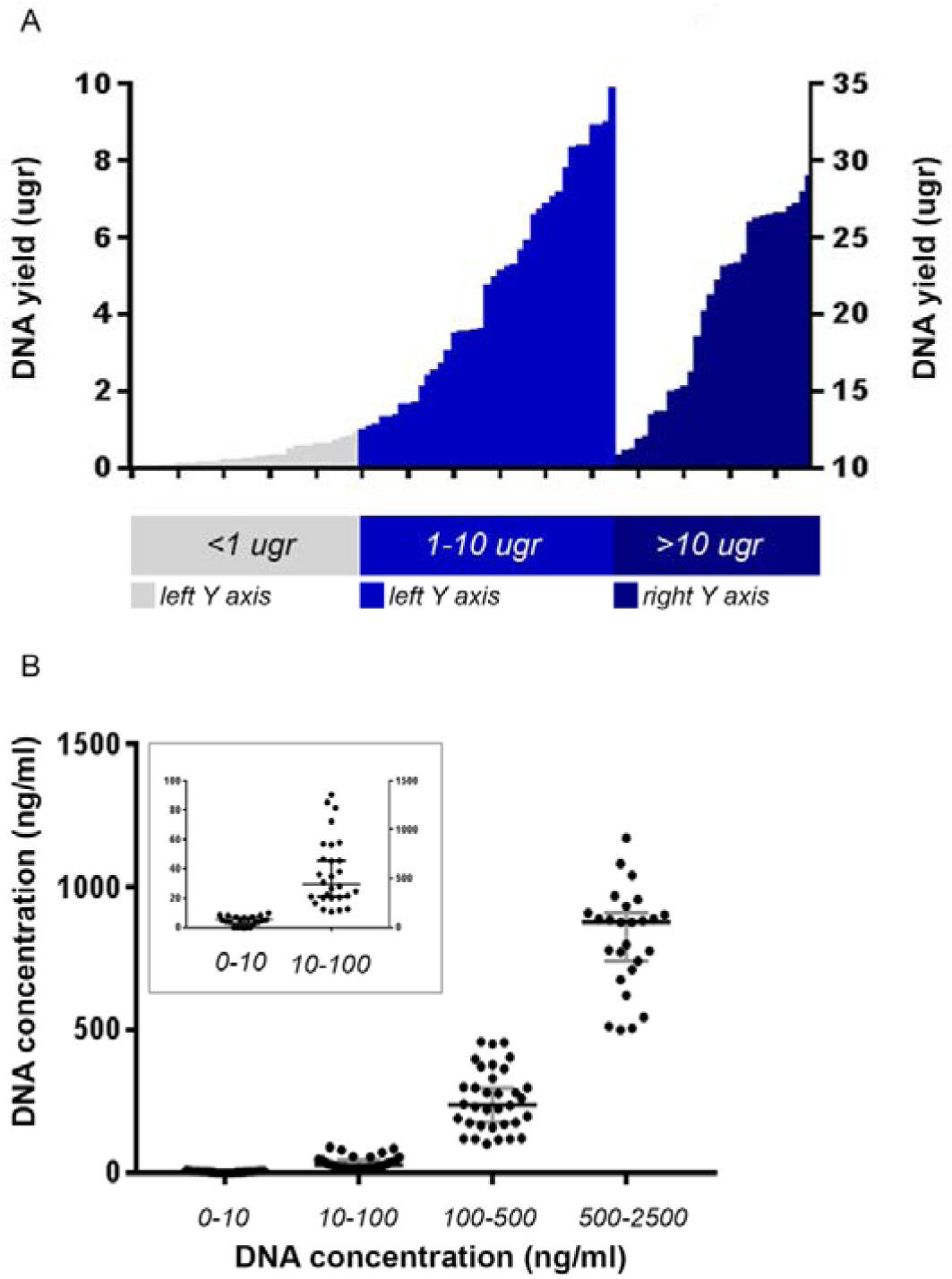
Maximum DNA yield of the whole analysed cohort from PROSPECT C and R patients. **A**) Cases with DNA yield >10ugr are plotted against right Y axis. **B**) cases divided according to their DNA concentration. Median value and with 95%CI represented in grey bars. In the small square cases with DNA concentration < 100 ng/ml are plotted against the left Y axis.

**Figure 3:**
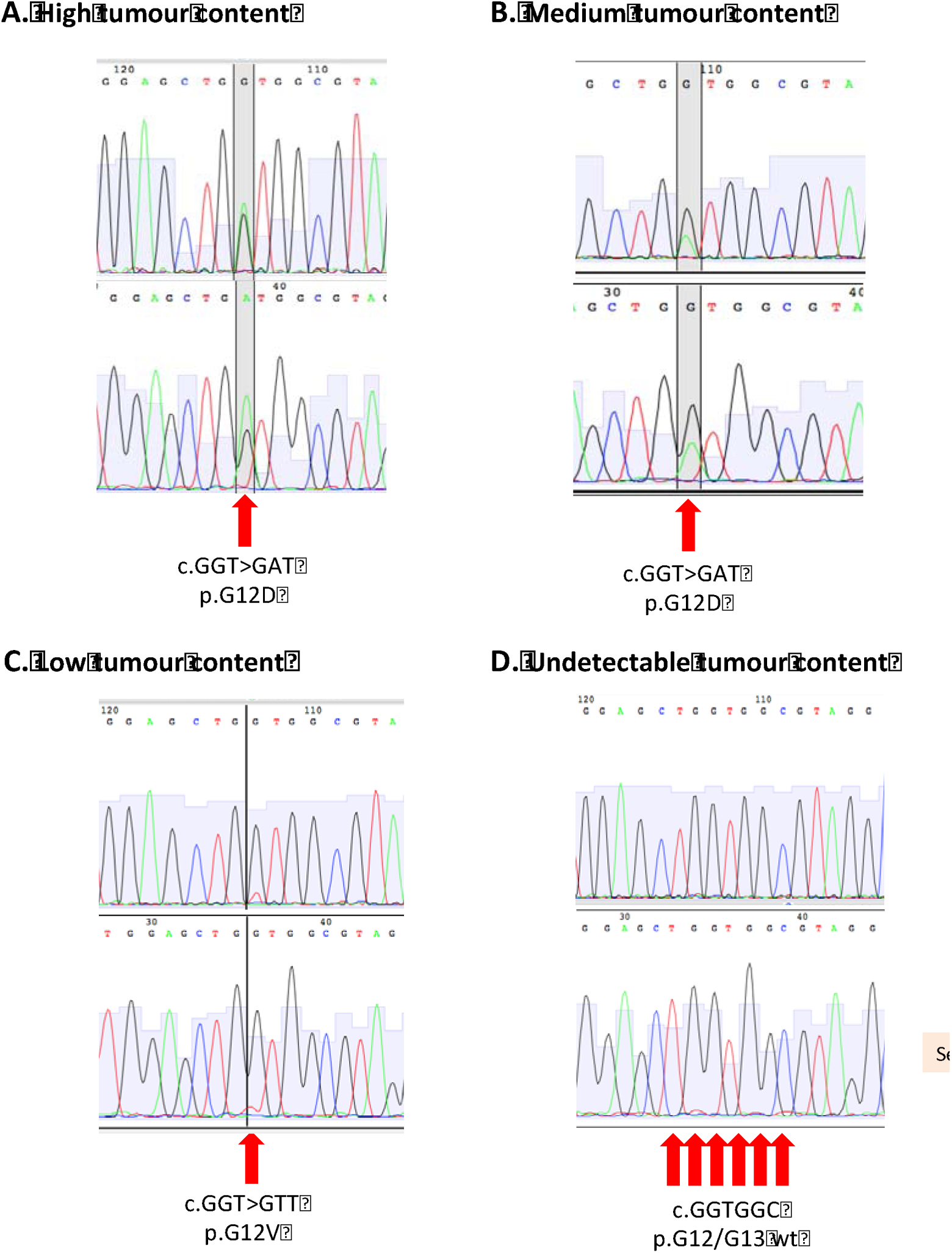
Example Sanger Sequencing traces for KRAS p.G12/G13.

## DISCUSSION

Tissue biopsies are often considered as the gold standard for diagnostic and research purposes; however, there are many logistical, technical and ethical challenges in the successful appliance of tissue biopsies in the clinic. To our knowledge, we present the largest dataset of tissue biopsies with a prospective validation cohort demonstrating high tumour yield and ability to perform genomic analysis via image-guided tissue sampling.

Biomarker discovery requires validation in prospective clinical trials; however, tissue collection procedures need to be optimised such that the valuable tissue obtained during trials is processed successfully [9, 10]. Moreover, even within a resource-friendly environment, molecular profiling studies have often suffered due to inadequacy of samples; failure rates reportedly vary between 15-33% [11-14]. Keeping these issues in view, we ensured that pre-biopsy scans were discussed in person with a radiologist and only the most amenable lesions were chosen for pre-treatment biopsies; experienced radiologists were then able to target multiple cores (6) from the periphery of the chosen lesions. The current study demonstrates that a strong infrastructure and good communication allows high quality tumour samples to be obtained in a time-efficient manner. The co-axial biopsy technique used at the Royal Marsden has the advantage of puncturing the capsule of solid organs (liver, kidney, spleen) only once, minimising the injury to normal tissues, improving patient experience and at the same time acquiring multiple large cores for diagnosis and molecular analysis. The application of pre-formed gelatin sponge sealing device at the biopsy tract provides a mechanical matrix that facilitates clotting. Gelfoam pledgets due their bulk, surface-acting hemostatic agents slow the flow of blood, protect the forming clot, and offer a framework for deposition of the cellular elements of blood decreasing the risk of major bleeding [15]. The grade III hemorrhage in our series was only 0.2%, which compares favourably with the 0.5%-2% seen in large series in the literature using tru-cut needles with or without co-axial technique [16, 17].

Common concerns about trials mandating research biopsies include the lack of patient understanding about the purpose of such studies, and the potential risks associated with additional interventional procedures within the research protocols [18-21]. In the case of our PROSPECT-C trial[1, 2], patients included could already access anti-EGFR antibody treatment via the cancer drug fund (CDF) independent of the research biopsy findings, which meant that the research biopsies were of no direct patient benefit. In order to ensure that patients clearly understood the purpose of their participation in PROSPECT-C and other research studies, a prospective patient based survey at the RM was performed. Remarkably, it showed that most patients who consented to a research biopsy gave an altruistic reason (e.g. to help research and/or others) as to why they agreed to participate [22]. A common concern regarding trial-related invasive intervention is procedure related complications. Notably, the biopsy complication rates in more than 500 patients in our cohort (including patients on PROSPECT studies) were extremely low and compared favourably with published literature [23, 24]. The technical reasons for success can be attributed to the use of large gauge co-axial needles, which enable multiple tissue cores to be sampled with a single pass. Subsequent application of gelatin foam pledgets via a co-axial cannula at withdrawal effectively seals the biopsy track and minimizes haemorrhage (<1%), thus enabling safe same-day discharge in the majority of patients. This technique however needs to be carefully considered in appropriate patients, for example, any attempt to biopsy lung paraenchyma would carry a significant risk of the gelfoam pledget deploying in a pulmonary vein resulting in systemic embolus.

Following the safe acquisition of biopsy material, the processing of tumour samples has its own challenges. Firstly, the acquired sample contains a mixture of cancer cells and stroma (connective tissue, blood vessels and inflammatory cells). It is well established that stromal infiltration may lead to problems in interpreting genomic data [25, 26]. In contemporaneous studies conducted at the RM (e.g. FOrMAT study), sample failure rates were high with only 16% of samples showing tumour content of more than 50% [27]. The FOrMAT study collected a range of GI tumour samples including pancreatic cancers, which are more likely to be dominated by inflammatory and stromal cells [28], but it relied on using only FFPE tissues. FFPE tissue has limitations for complex genomic studies as the DNA yield and quality is affected by the process of fixation and paraffin-embedding [29-33]. The PROSPECT studies benefited from parallel analysis using both FFPE and fresh frozen tissue, where the former was used for pathological assessment and the latter for molecular characterisation and genomic analysis. By utilising all available tissue cores as required, we achieved a gDNA extraction of >90% and an estimated tumour content of >20% in 87.27% of the cases. These data compare favourably with a recent large scale study comprising of >10,000 patients, who were subjected to a hybridization-based NGS panel capable of detecting all-protein coding mutations, copy number alterations, and selected promoter mutations and structural re-arrangements[34].

We next took into account the limitations of tumour estimates generated by subjective pathological assessment of tumour morphology and cellularity estimates. Cellularity can be estimated by quantifying the mutant alleles using technologies, such as Sanger or Ion Torrent sequencing, but this requires prior knowledge of the mutation [26, 35]; in PROSPECT-R Sanger sequencing was used to assess tumour content, as *RAS* mutation was a pre-requisite for entry into the study. However, only patients with no known *RAS* pathway mutation could participate in PROSPECT-C, so alternative techniques were required for tumour cellularity estimates. An unbiased statistical approach that directly measures tumour content from the DNA sample was therefore, which allowed us to take into account factors such as tumour ploidy and ITH.

## CONCLUSION

Oncologic management and clinical trial participation require accurate histological and molecular characterisation. Image-guided biopsies using large gauge co-axial needles enable multiple tissue cores to be obtained with a single pass. This increases the probability of diagnostic success for complex molecular analysis. Applying gelatin foam pledgets via the co-axial cannula following biopsy to seal the track reduces haemorrhagic risk and enables safe same day discharge in the majority of patients. By successfully obtaining sufficient number of tumour tissue samples within prospective trials, such studies can further understanding of tumour biology and help develop biomarkers of clinical and translational relevance. Ultimately, this will enhance the application of personalised medicine in the clinic.

## Data Availability

DNA and RNA sequencing data have been deposited in the European Genome Phenome short read archive and access can be obtained after signing a material transfer agreement which protects patient confidentiality and prohibits any attempts to re-identify patients.

## Funding

The authors acknowledge support from the National Institute for Health Research Biomedical Research Centre at The Royal Marsden NHS Foundation Trust and The Institute of Cancer Research.

## Conflict of interest

D.C. received research funding from: Roche, Amgen, Celgene, Sanofi, Merck Serono, Novartis, AstraZeneca, Bayer, Merrimack and MedImmune. I.C. has had advisory roles with Merck Serono, Roche, Sanofi Oncology, Bristol Myers Squibb, Eli-Lilly, Novartis, Gilead Science. He has received research funding from Merck-Serono, Novartis, Roche and Sanofi Oncology, and honoraria from Roche, Sanofi-Oncology, Eli-Lilly, Taiho. KK has advisory role with Bayer Oncology group. NV received honoraria from Merck Serono, Bayer and Eli-Lilly. All other authors declare no conflict of interest.

## Supplementary data

**Supplementary Figure 1:**
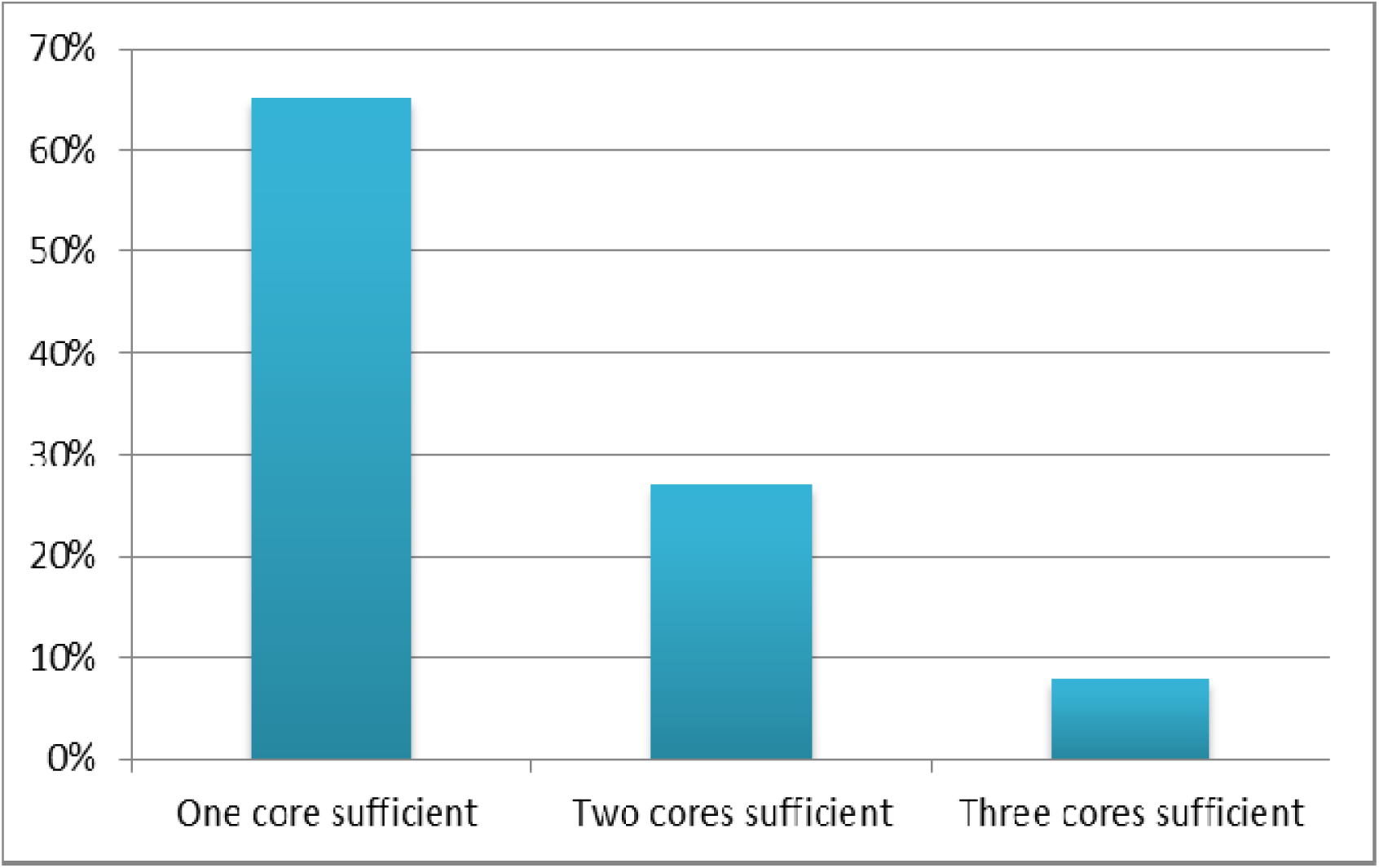
Average core requirement for sufficient gDNA extraction.

**Supplementary Table 1:**
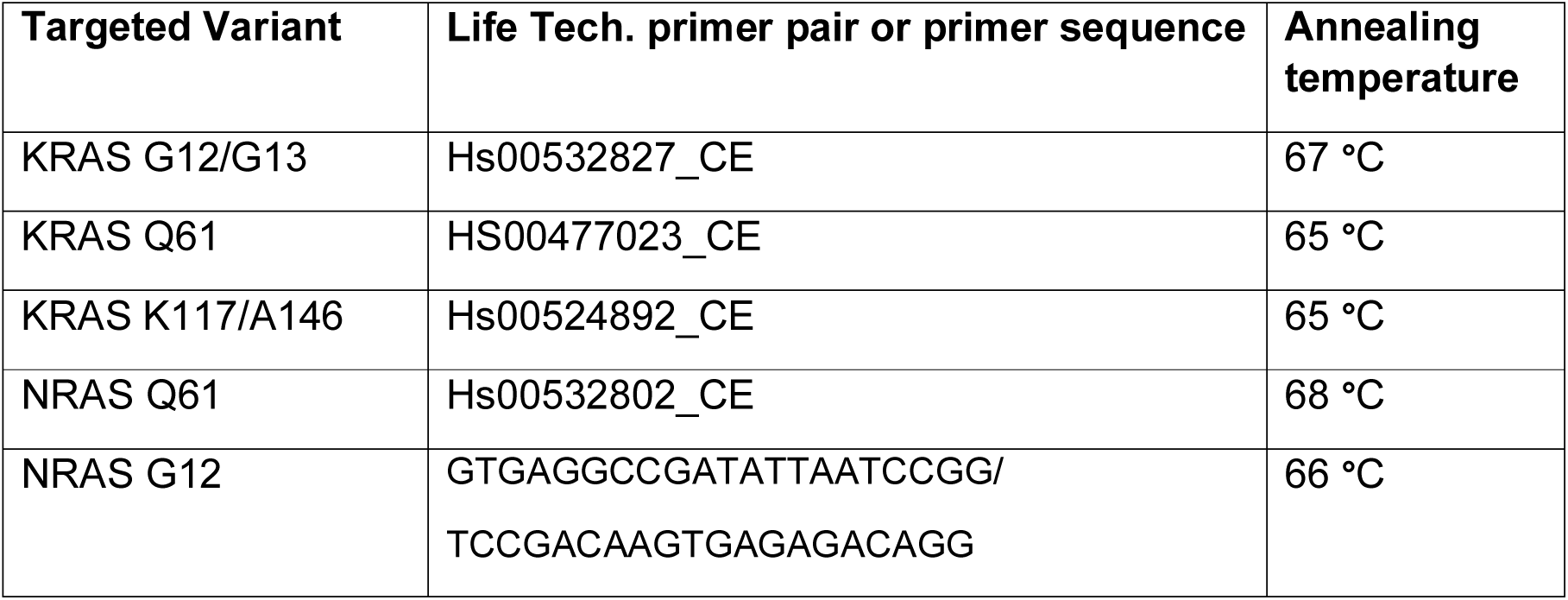
Sanger Sequencing Primers

**Supplementary Table 2:** Baseline demographics

| **Gender** | **N** | **%** |
| --- | --- | --- |
| Female | 237 | 51.86 |
| Male | 220 | 48.14 |
| Total | 457 | 100 |

| **Modality** | **N** | **%** |
| --- | --- | --- |
| USS | 290 | 63.46 |
| CT | 167 | 36.54 |
| Total | 457 | 100 |

| **Primary Tumour** | **Frequency** | **Percentage** |
| --- | --- | --- |
| Upper GI, CUP and hepatobiliary | 89 | 17.05 |
| lymphoproliferative disorders | 43 | 8.24 |
| urological | 60 | 11.49 |
| breast | 45 | 8.62 |
| gynaecological | 38 | 7.28 |
| lower GI | 144 | 27.59 |
| sarcoma | 49 | 9.39 |
| skin | 13 | 2.49 |
| thoracic | 40 | 7.66 |
| other (thyroid) | 1 | 0.19 |
| Total | 522 | 100 |

**Supplementary Table 3A:**
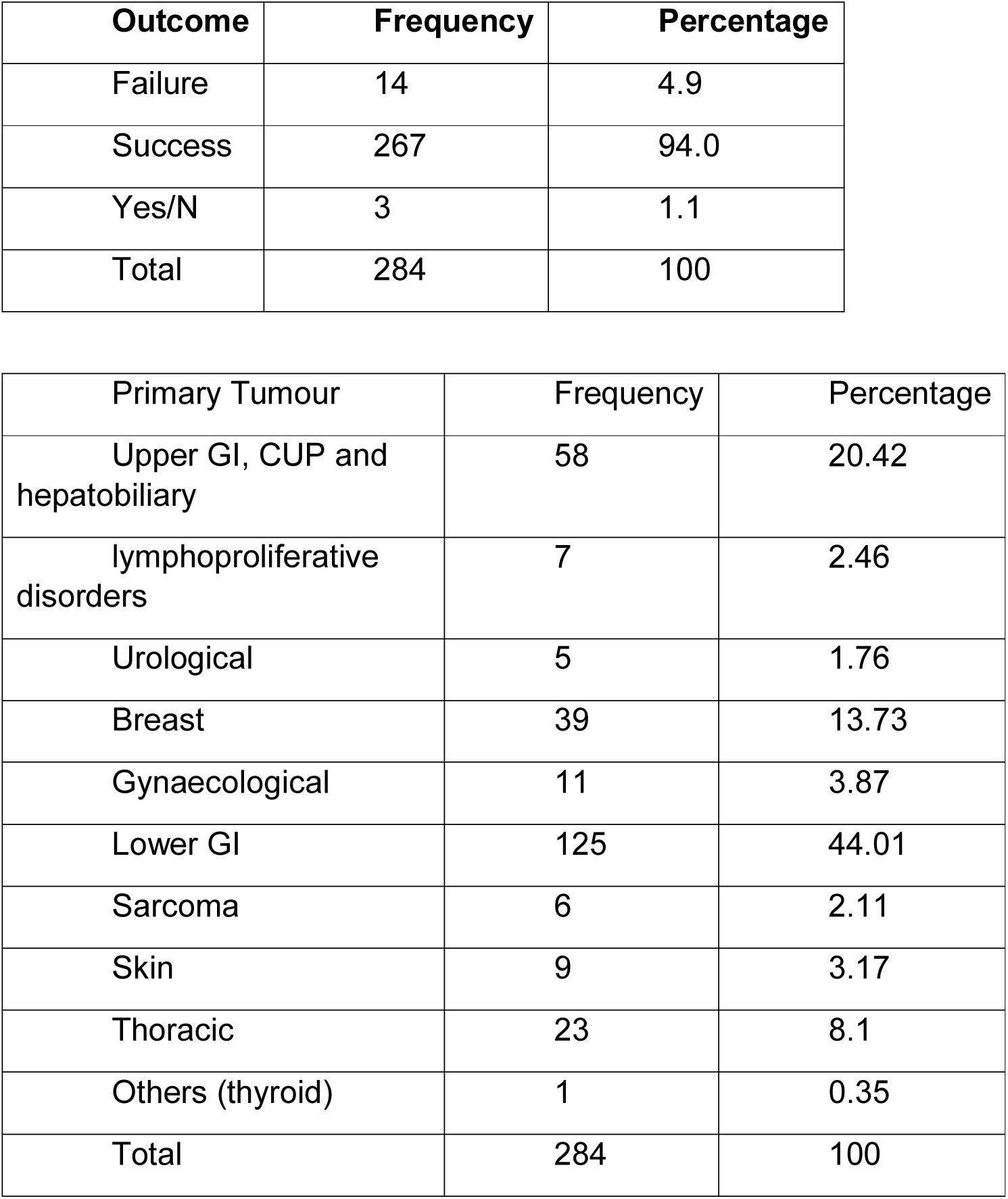
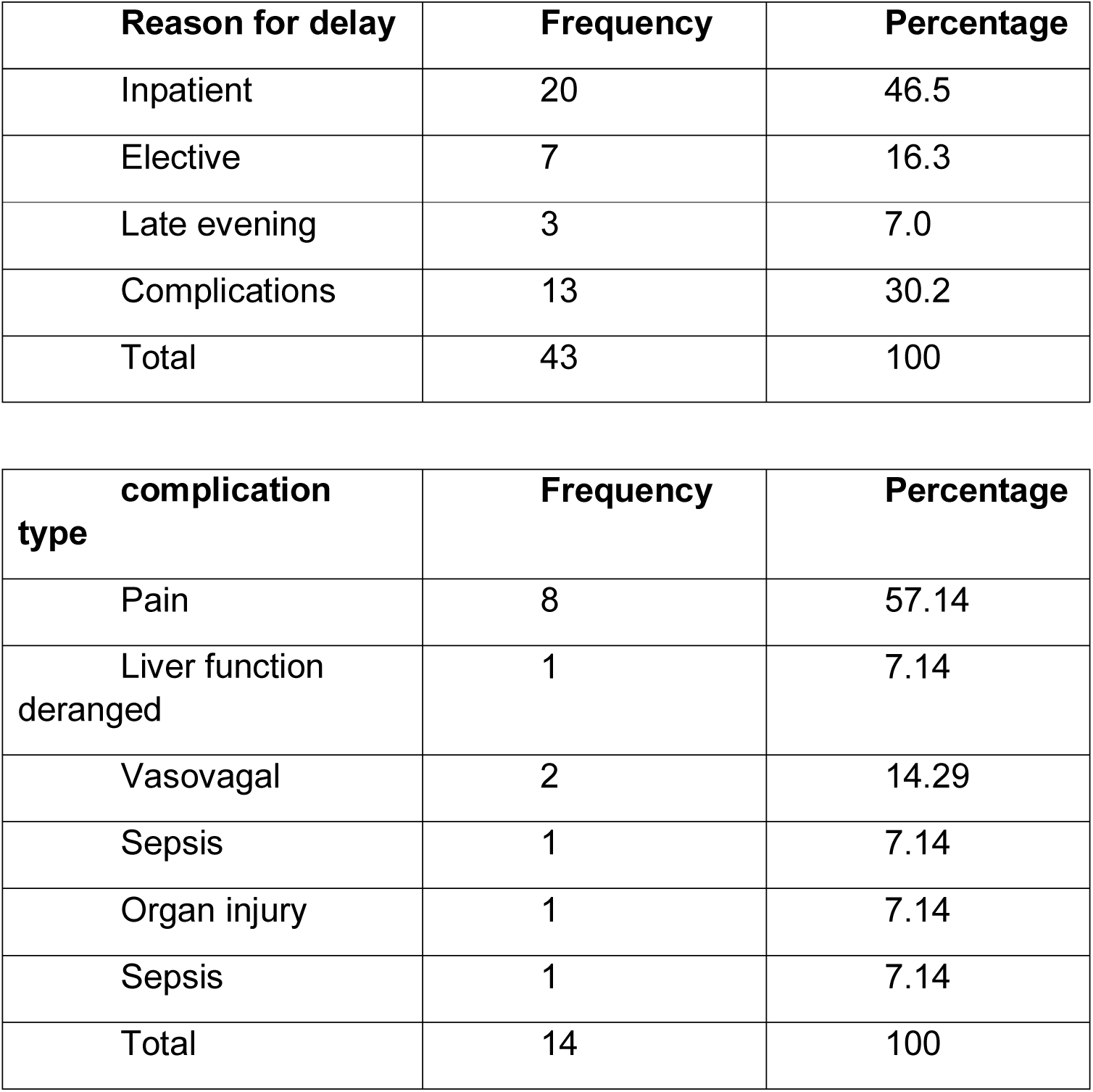

| **Outcome** | **Frequency** | **Percentage** |
| --- | --- | --- |
| Failure | 14 | 4.9 |
| Success | 267 | 94.0 |
| Yes/N | 3 | 1.1 |
| Total | 284 | 100 |

| **Primary Tumour** | **Frequency** | **Percentage** |
| --- | --- | --- |
| Upper GI, CUP and hepatobiliary | 58 | 20.42 |
| lymphoproliferative disorders | 7 | 2.46 |
| Urological | 5 | 1.76 |
| Breast | 39 | 13.73 |
| Gynaecological | 11 | 3.87 |
| Lower GI | 125 | 44.01 |
| Sarcoma | 6 | 2.11 |
| Skin | 9 | 3.17 |
| Thoracic | 23 | 8.1 |
| Others (thyroid) | 1 | 0.35 |
| Total | 284 | 100 |

| **Reason for delay** | **Frequency** | **Percentage** |
| --- | --- | --- |
| Inpatient | 20 | 46.5 |
| Elective | 7 | 16.3 |
| Late evening | 3 | 7.0 |
| Complications | 13 | 30.2 |
| Total | 43 | 100 |

| **complication**<br>**type** | **Frequency** | **Percentage** |
| --- | --- | --- |
| Pain | 8 | 57.14 |
| Liver function<br>deranged | 1 | 7.14 |
| Vasovagal | 2 | 14.29 |
| Sepsis | 1 | 7.14 |
| Organ injury | 1 | 7.14 |
| Sepsis | 1 | 7.14 |
| Total | 14 | 100 |

**Supplementary Table 3B:**

| **Outcome** | **Frequency** | **Percentage** |
| --- | --- | --- |
| Failure | 4 | 1.68 |
| Success | 234 | 98.32 |
| Total | 238 | 100 |

| **Site** | **Frequency** | **Percentage** |
| --- | --- | --- |
| Kidney | 35 | 14.71 |
| Abdominal | 44 | 18.49 |
| Pelvic | 60 | 25.21 |
| Lymph Node | 60 | 25.21 |
| Chest wall | 6 | 2.52 |
| Subcutaneous | 1 | 0.42 |
| Breast | 1 | 0.42 |
| Adrenal | 16 | 6.72 |
| Splenic | 5 | 2.1 |
| Pancreatic | 6 | 2.52 |
| Lung | 1 | 0.42 |
| Soft tissue | 2 | 0.84 |
| Omental/Peritoneal | 1 | 0.42 |
| Total | 238 | 100 |

| **Primary Tumour** | **Frequency** | **Percentage** |
| --- | --- | --- |
| Upper GI, CUP and hepatobiliary | 31 | 13.03 |
| Lymphoproliferative disorders | 36 | 15.13 |
| Urological | 55 | 23.11 |
| Breast | 6 | 2.52 |
| Gynaecological | 27 | 11.34 |
| Lower GI | 19 | 7.98 |
| Sarcoma | 43 | 18.07 |
| Skin | 4 | 1.68 |
| Thoracic | 17 | 7.14 |
| Total | 238 | 100 |

| **Delay** | **Frequency** | **Percentage** |
| --- | --- | --- |
| Inpatient | 15 | 42.86 |
| Elective | 10 | 28.57 |
| Precaution | 1 | 2.86 |
| Late evening | 5 | 14.29 |
| Complications | 4 | 11.43 |
| Total | 35 | 100 |

| **Complications** | **Frequency** | **Percentage** |
| --- | --- | --- |
| pain | 1 | 25 |
| hematuria | 1 | 25 |
| thrombosis | 1 | 25 |
| hemorrhage | 1 | 25 |
| Total | 4 | 100 |

**Supplementary Table 4:**

| Outcome | Liver | Other | Total |
| --- | --- | --- | --- |
| Failure | 14 | 4 | 18 |
|  | 4.98 | 1.68 | 3.47 |
| Success | 267 | 234 | 501 |
|  | 95.02 | 98.32 | 96.53 |
| Total | 281 | 238 | 519 |
|  | 100 | 100 | 100 |

**Association of clinical trial with outcome**
|  | Clinical trial |  |  |
| --- | --- | --- | --- |
| Outcome | no | yes | Total |
| Failure | 16 | 2 | 18 |
|  | 4.52 | 1.21 | 3.47 |
| Success | 338 | 163 | 501 |
|  | 95.48 | 98.79 | 96.53 |
| Total | 354 | 165 | 519 |
|  | 100 | 100 | 100 |

p: 0.07 from chi-squared test

## REFERENCES

[1] Khan K, Rata M, Cunningham D, Koh DM, Tunariu N, Hahne JC, et al. Functional imaging and circulating biomarkers of response to regorafenib in treatment-refractory metastatic colorectal cancer patients in a prospective phase II study. Gut. 2017.

[2] Khan KH, Cunningham D, Werner B, Vlachogiannis G, Spiteri I, Heide T, et al. Longitudinal Liquid Biopsy and Mathematical Modeling of Clonal Evolution Forecast Time to Treatment Failure in the PROSPECT-C Phase II Colorectal Cancer Clinical Trial. Cancer discovery. 2018.

[3] Cancer Genome Atlas Network. TCGA Tissue Sample Requirements: High Quality Requirements Yield High Quality Data. http://cancergenomenihgov/cancersselected/biospeccriteria. Accessed 22.05.16.

[4] Hsu MY, Pan KT, Chen CM, Lui KW, Chu SY, Lin YY, et al. CT-guided percutaneous core-needle biopsy of pancreatic masses: comparison of the standard mesenteric/retroperitoneal versus the trans-organ approaches. Clinical radiology. 2016;71:507–12.

[5] Olson MC, Atwell TD, Harmsen WS, Konrad A, King RL, Lin Y, et al. Safety and Accuracy of Percutaneous Image-Guided Core Biopsy of the Spleen. AJR American journal of roentgenology. 2016;206:655–9.

[6] Ocak S, Duplaquet F, Jamart J, Pirard L, Weynand B, Delos M, et al. Diagnostic Accuracy and Safety of CT-Guided Percutaneous Transthoracic Needle Biopsies: 14-Gauge versus 22-Gauge Needles. Journal of vascular and interventional radiology: JVIR. 2016;27:674–81.

[7] Vlachogiannis G, Hedayat S, Vatsiou A, Jamin Y, Fernandez-Mateos J, Khan K, et al. Patient-derived organoids model treatment response of metastatic gastrointestinal cancers. Science (New York, NY). 2018;359:920–6.

[8] Brannon AR, Vakiani E, Sylvester BE, Scott SN, McDermott G, Shah RH, et al. Comparative sequencing analysis reveals high genomic concordance between matched primary and metastatic colorectal cancer lesions. Genome biology. 2014;15:454.

[9] Diamandis M, White NM, Yousef GM. Personalized medicine: marking a new epoch in cancer patient management. Molecular cancer research: MCR. 2010;8:1175–87.

[10] Wistuba, II, Gelovani JG, Jacoby JJ, Davis SE, Herbst RS. Methodological and practical challenges for personalized cancer therapies. Nature reviews Clinical oncology. 2011;8:135–41.

[11] Ferté C, Massard C, Ileana E, Hollebecque A, Lacroix L, Ammari S, et al. Abstract CT240: Molecular screening for cancer treatment optimization (MOSCATO 01): a prospective molecular triage trial; Interim analysis of 420 patients Cancer research. 2014;74; CT2140.

[12] Andre F, Bachelot T, Commo F, Campone M, Arnedos M, Dieras V, et al. Comparative genomic hybridisation array and DNA sequencing to direct treatment of metastatic breast cancer: a multicentre, prospective trial (SAFIR01/UNICANCER). The lancet oncology. 2014;15:267–74.

[13] Meric-Bernstam F, Brusco L, Shaw K, Horombe C, Kopetz S, Davies MA, et al. Feasibility of Large-Scale Genomic Testing to Facilitate Enrollment Onto Genomically Matched Clinical Trials. Journal of clinical oncology: official journal of the American Society of Clinical Oncology. 2015;33:2753–62.

[14] Le Tourneau C, Delord JP, Goncalves A, Gavoille C, Dubot C, Isambert N, et al. Molecularly targeted therapy based on tumour molecular profiling versus conventional therapy for advanced cancer (SHIVA): a multicentre, open-label, proof-of-concept, randomised, controlled phase 2 trial. The lancet oncology. 2015;16:1324–34.

[15] Lungren MP, Lindquester WS, Seidel FG, Kothary N, Monroe EJ, Shivaram G, et al. Ultrasound-Guided Liver Biopsy With Gelatin Sponge Pledget Tract Embolization in Infants Weighing Less Than 10 kg. Journal of pediatric gastroenterology and nutrition. 2016;63:e147–e51.

[16] Atwell TD, Smith RL, Hesley GK, Callstrom MR, Schleck CD, Harmsen WS, et al. Incidence of bleeding after 15,181 percutaneous biopsies and the role of aspirin. AJR American journal of roentgenology. 2010;194:784–9.

[17] Lipnik AJ, Brown DB. Image-Guided Percutaneous Abdominal Mass Biopsy: Technical and Clinical Considerations. Radiologic clinics of North America. 2015;53:1049–59.

[18] Pentz RD, Harvey RD, White M, Farmer ZL, Dashevskaya O, Chen Z, et al. Research biopsies in phase I studies: views and perspectives of participants and investigators. Irb. 2012;34:1–8.

[19] Olson EM, Lin NU, Krop IE, Winer EP. The ethical use of mandatory research biopsies. Nature reviews Clinical oncology. 2011;8:620–5.

[20] Saggese M, Dua D, Simmons E, Lemech C, Arkenau H. Research biopsies in the context of early phase oncology studies:clinical and ethical considerations. Oncology Reviews. 2013;7.

[21] Peppercorn J, Shapira I, Collyar D, Deshields T, Lin N, Krop I, et al. Ethics of mandatory research biopsy for correlative end points within clinical trials in oncology. Journal of clinical oncology: official journal of the American Society of Clinical Oncology. 2010;28:2635–40.

[22] Moorcraft SY, Begum R, Cunningham D, Peckitt C, Baratelli C, Gillbanks A, et al. Attitudes of Patients With Gastrointestinal Cancers Toward Research Biopsies. Clin Colorectal Cancer. 2016.

[23] Tacher V, Le Deley MC, Hollebecque A, Deschamps F, Vielh P, Hakime A, et al. Factors associated with success of image-guided tumour biopsies: Results from a prospective molecular triage study (MOSCATO-01). Eur J Cancer. 2016;59:79–89.

[24] Howlett DC, Drinkwater KJ, Lawrence D, Barter S, Nicholson T. Findings of the UK national audit evaluating image-guided or image-assisted liver biopsy. Part II. Minor and major complications and procedure-related mortality. Radiology. 2013;266:226–35.

[25] Laird PW. Principles and challenges of genomewide DNA methylation analysis. Nat Rev Genet. 2010;11:191–203.

[26] Song S, Nones K, Miller D, Harliwong I, Kassahn KS, Pinese M, et al. qpure: A tool to estimate tumor cellularity from genome-wide single-nucleotide polymorphism profiles. PLoS One. 2012;7:e45835.

[27] Moorcraft SY, Gonzalez de Castro D, Cunningham D, Jones T, Walker BA, Peckitt C, et al. Investigating the feasibility of tumour molecular profiling in gastrointestinal malignancies in routine clinical practice. Annals of oncology: official journal of the European Society for Medical Oncology / ESMO. 2018;29:230–6.

[28] Erkan M, Reiser-Erkan C, Michalski CW, Kleeff J. Tumor microenvironment and progression of pancreatic cancer. Experimental oncology. 2010;32:128–31.

[29] Koshiba M, Ogawa K, Hamazaki S, Sugiyama T, Ogawa O, Kitajima T. The effect of formalin fixation on DNA and the extraction of high-molecular-weight DNA from fixed and embedded tissues. Pathol Res Pract. 1993;189:66–72.

[30] Howat WJ, Wilson BA. Tissue fixation and the effect of molecular fixatives on downstream staining procedures. Methods. 2014;70:12–9.

[31] Zsikla V, Baumann M, Cathomas G. Effect of buffered formalin on amplification of DNA from paraffin wax embedded small biopsies using real-time PCR. Journal of clinical pathology. 2004;57:654–6.

[32] Cree IA, Deans Z, Ligtenberg MJ, Normanno N, Edsjo A, Rouleau E, et al. Guidance for laboratories performing molecular pathology for cancer patients. Journal of clinical pathology. 2014;67:923–31.

[33] Cancer Research UK. CRUK Stratified Medicine Program. http://wwwcancerresearchukorg/sites/default/files/smp1_booklet_12_-_no_markspdf Accessed 140516.

[34] Zehir A, Benayed R, Shah RH, Syed A, Middha S, Kim HR, et al. Mutational landscape of metastatic cancer revealed from prospective clinical sequencing of 10,000 patients. Nature medicine. 2017;23:703–13.

[35] Rothberg JM, Hinz W, Rearick TM, Schultz J, Mileski W, Davey M, et al. An integrated semiconductor device enabling non-optical genome sequencing. Nature. 2011;475:348–52.

